# Using evidence-based strategies, including coverage of remote cardiac rehabilitation, to increase cardiac rehabilitation participation - a community health insurance plan’s experience

**DOI:** 10.1101/2024.11.07.24316943

**Authors:** Zainab Magdon-Ismail, Sheree Murphy, Vickie Gauthier, Tiana Wyrick, Cori Rowe, Rachael Austin, Pennie Cuevas, Jeremy Pickreign, Bruce Coplin

## Abstract

**Background:** Barriers to referral, enrollment, and participation in cardiac rehabilitation (CR) contribute to low rates of completion despite known benefits. Barriers are system, provider and patient related. In this observational cohort quality improvement study, we examined the impact a community-based, not-for-profit health insurance plan had on barriers to CR participation.

**Methods:** The Capital District Physicians’ Health Plan (CDPHP) in Albany, New York developed and implemented a cardiac rehabilitation initiative (CRI) to increase CR rates using evidence-based strategies. CDPHP: 1) eliminated patient cost-share, 2) covered remote CR (RCR), 3) implemented physician valued-based incentives, 4) presented metrics to providers, 5) educated providers and patients, and 6) dedicated staff to facilitating enrollment. Chi-square tests were used to identify differences among patients who enrolled in facility-based CR (FBCR), RCR and no CR. CR enrollment rate distributions were evaluated between Q1, 2021 and Q2, 2022.

**Results:** A total of 1,736 patients with varying cardiac conditions were eligible for CR in the study period. Between Q1, 2021 and Q2, 2022, enrollment went from 11.1% (32/286) to 16.2% (50/308) in FBCR; 0.7% (2/286) to 10.7% (33/308) in RCR; and 11.9% (34/286) to 26.9% (83/308) overall (*P*<.0001). Time to enrollment went from 40 to 47 days for FBCR (P=0.1792), 53 to 20 days for RCR (*P*<.0001) and 43 to 36 days overall (*P*=0.3348). Older patients were more likely to enroll in CR as were patients who underwent cardiac procedures.

**Conclusions:** The CRI created a call-to-action among providers to address CR referral and enrollment. RCR increased CR rates and were additive to FBCR rates, suggesting that the introduction of RCR will not displace FBCR. Time to enrollment improved overall, driven by improvement in those enrolling in RCR. Increasing CR engagement requires coordinated effort from stakeholders—cardiology providers, hospitals, CR providers and health plans.

## Background

Despite its American Heart Association/American College of Cardiology (AHA/ACC) class 1a recommendation, participation in cardiac rehabilitation (CR) across the United States (US) remains low.^1–4^ Only a quarter of patients engage in CR and only a quarter of those complete it.^1–4^ A 2022 study of Medicare beneficiaries in 2017 revealed that only 28.6% of patients completed ≥ 1 session within a year of discharge from a qualifying event and of those, only 27.6% completed all 36 sessions.^4^ New York State (NYS) ranked second to last nationally in the study with only 15.6% of patients completing ≥ 1 session. Million Hearts, a national initiative jointly led by the Centers for Disease Control and Prevention (CDC) and the Centers for Medicare and Medicaid Services (CMS) has set a goal to increase CR participation to 70%.^5^

The evidence-based benefits of CR attributed to fitness and diet/nutrition as well as physiologic effects to the body have shown a 13% to 24% reduction in total mortality over 1 to 3 years, as well as a 31% decrease in rehospitalizations over one year.^6–11^ Additionally, patients experience an increase in their physical function and overall quality of life. ^6–10^ Common barriers to patients attending CR are lack of convenient options for where to receive rehab, travel, work, and co-pays.^1–10, 12^. Because the benefits of CR are dose related, adherence and completion of the regimen are key.^14–16^ While CR is indicated for most cardiac patients, often those with more acute disease get referred and enroll, while those with chronic conditions, such as heart failure, are less likely to be considered by providers.^1–4,13^

COVID-19 created a need for alternate CR delivery models.^17^ While remote cardiac rehabilitation (RCR) solutions have been available for decades and show similar results to facility-based cardiac rehabilitation (FBCR) in eligible patients, there has been a call to evaluate their effectiveness in varied health systems and amongst diverse populations, as most studies have been conducted within the Kaiser and Veterans Affairs (VA) systems. ^17–23^

A thorough search of the literature revealed no studies in the US of a non-integrated health plan addressing CR participation, including but not limited to offering RCR. The focus of this observational cohort study was to examine the introduction of RCR as well as the referral, enrollment and participation rates for CR in a non-integrated health system following quality improvement efforts introduced by a health insurance plan.

## Methods

This study followed the STrengthening the Reporting of OBservational studies in Epidemiology (STROBE) and Standards for QUality Improvement Reporting Excellence (SQUIRE) reporting guidelines. The Sterling Institutional Review Board reviewed this initiative; patient informed consent was not required.^24^

### Context

The Capital District Physicians’ Health Plan (CDPHP) is a physician-founded, not-for-profit community health insurance plan serving approximately 40% of 1.1 million people in the Capital Region of New York State.^25^ Plan membership consists of commercial (60%), Medicare (10%), Medicaid (20%) and as well as administrative services (ASO) (10%) members. At the time of the study, CDPHP was a non-integrated health plan.

In 2019, CDPHP examined the use of CR among key hospitals in its coverage area and determined that only 16% of members had at least one CR visit, with 15% of those completing all their CR sessions. Based on these findings, CDPHP planned to identify and address barriers to referral, enrollment and adherence to CR and launch a cardiac rehabilitation initiative (CRI).

Between October 2019 and June 2022, CDPHP reviewed and implemented evidence-based strategies to increase referral, enrollment, and adherence to CR noted in the literature and outlined in the CDC’s Cardiac Rehabilitation Change Package which details strategies to increase enrollment, participation and adherence in CR working with various stakeholders (Table 1).^26–33^ CDPHP created a multidisciplinary team of health plan physicians, actuarial staff, care managers and quality improvement staff in conjunction with external partners, including the New York State Department of Health (NYS DOH). The following efforts, described in detail below, were implemented 1) elimination of patient cost-share (co-pays/co-insurance), 2) coverage of RCR, 3) valued based incentives to physicians based on patient education about CR and patient completion of CR, 4) focus on metrics, specifically the American Heart Association/American College Cardiology Clinical Performance and Quality Metrics for Cardiac Rehabilitation^34^ and the National Committee on Quality Assurance (NCQA) Healthcare Effectiveness Data and Information Set (HEDIS) metrics related to CR,^35^ 5) provider and patient engagement efforts around CR and 6) dedicated staff to identifying and facilitating enrollment in CR, including review of gap lists. The official start of the CRI offering was April 1^st^ 2021.

**Table 1:** Capital District Physicians’ Health Plan Initiatives in Comparison to the Millions Heart Cardiac Rehab Change Package^26^.

| **System Change** |  |  |  |
| --- | --- | --- | --- |
| **Capital District Physicians' Health Plan (CDPHP) Initiative** | **Change Concept** | **Change Ideas** | **CDPHP Implementation/Tools & Resources** |
| 1. Elimination of Co-Pays | Reduce Cost Sharing Barriers for cardiac rehabilitation (CR) Services | Assist patients with high out-of-pocket costs or economic burden to navigate payment options | Eliminated co-pays where possible |
| 2. Coverage of Remote Cardiac Rehabilitation (RCR) | Develop Flexible Models That Better Accommodate Patient Needs | Develop hybrid model of home-based and facility-based program that includes key components of CR | Provided RCR option<br>Provided option for members to participate in both facility-based cardiac rehabilitation (FBCR) & RCR (hybrid) |
| 3. Valued-based Incentives to physicians | Use Clinician Follow-up to Bolster Enrollment or Participation | Engage referring clinicians by providing process reports and completion of program outcomes | Incented cardiology practices (monetarily) to educate patients on the importance of CR and also incented cardiology practices for each patient who completed CR |
| 4. Focus on metrics, specifically HEDIS metrics related to CR | Make CR a Health System Priority | Use CR referral enrollment and participation as quality-of-care indicators | Used HEDIS data to inform implementation activities through the use of data dashboards |
|  | Use Data to Drive Improvements in Referrals to CR | Use performance measures in a quality improvement system<br>Regularly provide dashboard with CR referral metrics, goals, and performance | Performance on HEDIS metrics was shared with cardiology practices and hospitals |
|  | Reduce delay from discharge to 1<sup>st</sup> CR Appointment | Before hospital discharge establish an early, within 12 days of discharge, outpatient appointment. | Worked with all CR providers on reducing time from discharge to first appointment<br>Implemented CR awareness initiative to help cardiology offices and hospitals educate and refer patients to CR |
|  | Use Clinician Follow-up to Bolster Enrollment or Participation | Engage referring clinicians by providing letters that highlight non-enrolled patients for clinician follow-up<br>Engage referring clinicians by providing progress reports and completion of program outcomes | Provided gap list to large provider groups |
| 5. Community Education regarding CR to cardiology practices, hospitals, cardiac rehab providers and patients | Educate Patients About the Benefits of Outpatient CR | Use videos to describe your CR program and the impact of CR on health outcomes before hospital discharge or at the beginning of outpatient CR | Developed an educational video on the importance of CR and what a patient's options are. Video could be used in all settings as well as sent to patients for review outside of a healthcare setting. |
|  | Educate Patients About the Benefits of Outpatient CR | Provide patient education materials that convey CR benefits | Developed education materials for inpatient and outpatient use. |
| 6. Dedicated staff to identifying and facilitating enrollment in CR | Make CR a Health System Priority | Establish a hospital champion, such as a quality of care leader or CR administrator | Established the following:<br>-physician champion<br>-implementation champion<br>-implementation team (inpatient and outpatient nurse case managers, social workers) |
|  | Make CR a Health System Priority | Engage care team in CR to ensure their buy-in in CR | Inpatient and outpatient care managers worked with providers in cardiology practices and hospitals to improve referral to CR workflows<br>Education (via webinar and in person) was provided to providers in cardiology practices and hospitals |
|  | Incorporate Referral into Standardized Process of Care | Develop a standard process for informing an external CR program of a referred patient<br>Implement standardized paper/faxed referral to CR from an inpatient setting | Worked with virtual CR provider through a portal and with regular touch base meetings to ensure that referred patients were captured |
|  | Incorporate Referral into Standardized Process of Care | Develop and communicate a standardized referral process or policy for patients | Worked with virtual CR provider through a portal and with regular touch base meetings to ensure that referred patients were captured. |
|  | Use Data to Drive Improvement in Referrals to CR | Identify patients who had a cardiac event without a referral to a CR program | Reviewed gap lists monthly. |
|  | Educate Patients about the Benefits of Outpatient CR | Deploy an inpatient “liaison” to help educate, refer, schedule, and enroll eligible patients in outpatient CR | Inpatient care managers educated, referred patients to CR.<br>Members interested in CR sent to CR care manager point team |
|  | Identify Populations at Risk for Low Engagement | Know the characteristics that are predictive of attendance and drop-out to identify patients at particular risk, to offer extra support | Worked with RCR provider who reached out to CDPHP CR liaison to assist with patients who were: unable to be reached, were unwilling to participate, and who had low engagement |
|  | Improve Patient Engagement | Incorporate motivational and financial incentives for meeting goals for session attendance | CDPHP reached out to patients if notified of a member’s unwillingness to participate or low engagement |
|  | Incorporate Referral into Standardized Processes of Care for Eligible Patients | Include referral to CR in order sets for appropriate patients; Incorporate into EHR as appropriate | Worked with providers to explore use of EHR tools to automate referrals. Also explored and incorporated, to the extent possible, automation capabilities to flag health plan team members to refer a patient to CR |

\*Eliminated patient cost share for all groups (commercial, Medicare, Medicaid) but those on high deductible plans.

In September of 2020, CDPHP partnered with the NYS DOH/Health Research Inc. (HRI) to support the implementation and evaluation of the CRI initiative. The project was part of NYS DOH/HRI’s Diabetes and Heart Disease and Control program funded by the CDC (DP10-1815).^36^

#### 1. Elimination of Patient Cost Share

Internal review of CR usage and a cost-benefit analysis informed the decision to eliminate patient cost-share (co-pays/co-insurance) where possible. Some plans such as high deductible plans and plans where CDPHP was contracted for administrative services only did not allow for cost-share waiver without certain conditions being met. CDPHP filed intent to waive member cost-share with the Center for Medicare and Medicaid Services (CMS) for Medicare patients, and the New York State Department of Financial Services (DFS) for commercial offerings. In this cohort study approximately 85% of patients did not have a cost share.

#### 2. Coverage of Remote Cardiac Rehabilitation (RCR)

CDPHP contracted with Movn Health a national RCR provider to deliver CR. The Movn program is based on the MULTIFIT model ^21–22, 37^ and patients participating received a cellular enabled weight scale and blood pressure cuff, a smart watch (Apple Watch or Garmin), exercise resistance bands, education material as well as access to a phone app to track their progress and communicate with their care manager. Patients had initial intake visits and were set up with an exercise regimen, counseling and one-on-one check-ins for 12 weeks followed by monitoring for 9 months. Members were given the option to switch between FBCR and RCR once during the course of CR. Movn was set up ideally for patients in the low to moderate risk categories based on the American Association of Cardiovascular and Pulmonary Rehabilitation (AACVPR) stratification, though exceptions for higher risk patients could be made with physician approval.^38^

#### 3. Value Based Incentives to Physicians

To drive improvement in referrals to CR, CDPHP developed three synergistic provider incentives: educating patients on the importance of CR, facilitating patient enrollment, and encouraging program completion. The incentives were offered to cardiology practices participating in CDPHP’s larger quality and specialty value-based programs. In addition to negotiated payments, practices who met certain CR performance benchmarks were eligible to receive additional incentive payments.

#### 4. Focus on Metrics

To track and monitor improvements in enrollment, participation, and completion of CR, CDPHP adopted 6 key metrics which were a blend of the AHA/ACC Cardiology Clinical Performance and Quality Metrics for Cardiac Rehabilitation^34^ and HEDIS Cardiac Rehabilitation Engagement (CRE) metrics ^35^: 1. Enrollment in CR (first visit)^34^, 2. Time to enrollment in CR (from qualifying event to first visit—goal 21 days)^34^, 3. Initiation (>2 visits)^35^, 4. Engagement 1 (> 12 visits)^35^, 5. Engagement 2 (> 24 visits)^35^, and 6. Achievement (>36 visits)^34, 35^. Data was shared with providers to show performance relative to their peers on a quarterly basis.

#### 5. Provider & Patient Engagement

An educational campaign was rolled out on the CRI Program to hospital systems, cardiology practices (including cardiac surgery practices), FBCR programs, and patients. Patients were educated via patient newsletters, blogs and on the CDPHP website. A patient education video outlining the benefits of CR and the options of FBCR and RCR was developed to be used in hospitals and cardiology practices (see Appendix A). All providers received individualized education in person or via or web conference. Providers were encouraged to provide eligible patients the option to attend FBCR or RCR

CDPHP established frameworks to assess CR referral, enrollment and participation across providers and patients of differing types. Review of these frameworks, in turn, shaped the content for monthly follow-up visits with all providers which included sharing of baseline data, assessing current referral processes and setting forth progressive next steps. Detailed workflows and tools used with providers can be found in Appendices B-D.

For the initial months of program deployment, a patient satisfaction survey was administered over the phone to help understand referral patterns as well as ensure that patients were equally satisfied with the FBCR and RCR (see Appendix E).

#### 6. Staff for Identification and Facilitation of Enrollment in CR

CDPHP established a physician administrative champion to coordinate the rollout of the CRI. A project lead was established to educate all inpatient care managers (CDPHP staff located within highest volume hospitals) and outpatient care managers (staff who managed patients post hospital discharge) on how to identify patients eligible for CR, speak to the importance of CR and facilitate enrollment in either FBCR or RCR based on member eligibility and preference. Additionally, the project lead coordinated referrals if needed, monitored daily admission/discharge feeds and reviewed gap lists monthly to ensure that members were being identified and referred to CR.

### Study Population

To study the effect of the quality improvement strategies, CDPHP adopted the afore mentioned process and outcome measures. Data were collected on 1,762 CDPHP members ≥18 years, discharged from January 1^st^ 2021-June 30^th^ 2022 with the following diagnoses: myocardial infarction (MI), percutaneous coronary intervention (PCI), valve procedure (Valve), coronary artery bypass graft (CABG), transplant or a combination of these procedures. CDPHP members who were Medicare, Medicaid and commercial were included. Since the population of interest were those considered for HEDIS measures, patients with heart failure were excluded.

### Statistical Analyses

We examined the sociodemographic and clinical characteristics of the cohort using means (standard deviation) and medians (interquartile range) for continuous variables and proportions for dichotomous variables. Of the five monitored metrics, we assessed 1) enrollment in CR and 2) time to enrollment in CR as they related to program uptake, as opposed to follow through, and these measures were readily available at the time of manuscript preparation. Chi-square was employed to assess distributional differences among patient characteristics stratified by type of CR, and to assess differences in participation in CR over time. A trend line was fit to the time to enrollment to assess whether the trend over time was significant.

SAS Enterprise Guide version 7.15 (SAS Institute, Inc., Cary, NC) was used for all analyses. *P*-value <0.05 was considered statistically significant.

## Results

### CRI Development & Implementation

Table 2 displays the set up and implementation timeline. CRI development and implementation took a total of 18 months from program conception to program implementation (October 2019-March 2021). Filing changes for cost share waivers with the governing bodies for Medicaid, Medicare and commercial took 9 months. Contracting with Movn Health took 4 months.

**Table 2:**
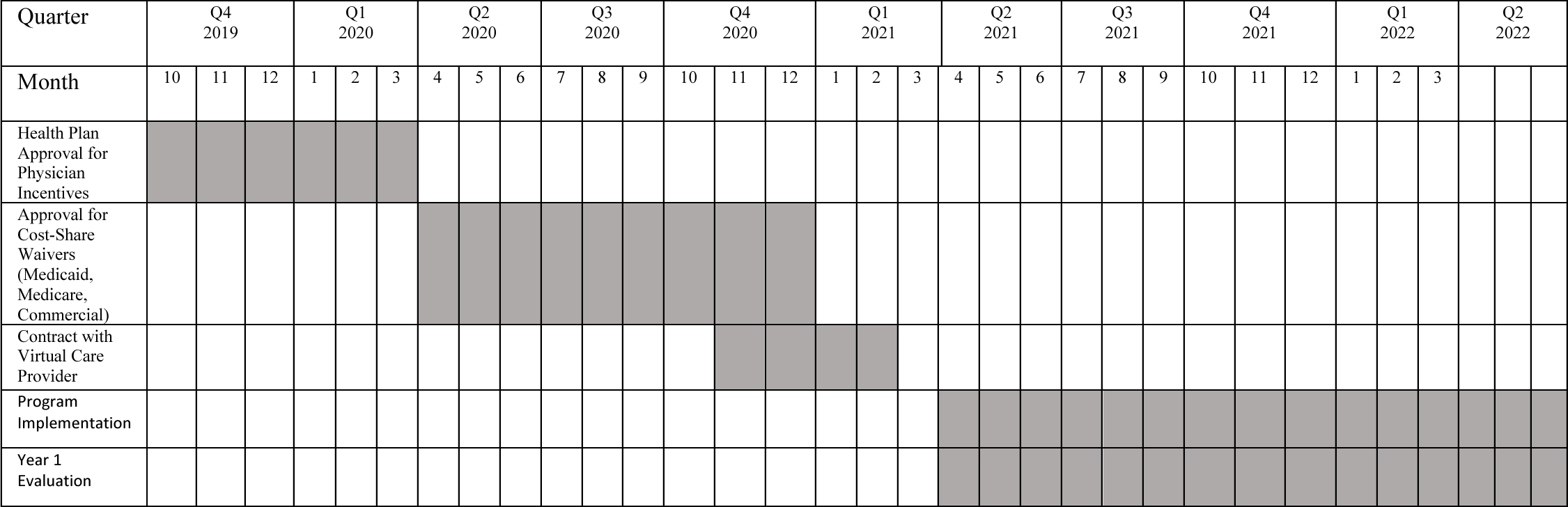
Cardiac Rehab Initiative Development and Implementation Timeline.

### Patient Demographic Characteristics

Table 3 details all the patient sociodemographic and clinical characteristics of the 1,762 patients included in the cohort. The mean age was 61.6 ± 11.7 with the majority of patients (82.3%) falling between the ages of 50 and 80 years old. Males accounted for 68.0% of the cohort. The top three qualifying diagnoses were MI (33.5%), PCI (24.2%), and MI PCI (18.7%). Patient’s insurance coverage was as follows: Medicare (34.2%), Medicaid (24.1%), commercial (41.6%).

**Table 3:**
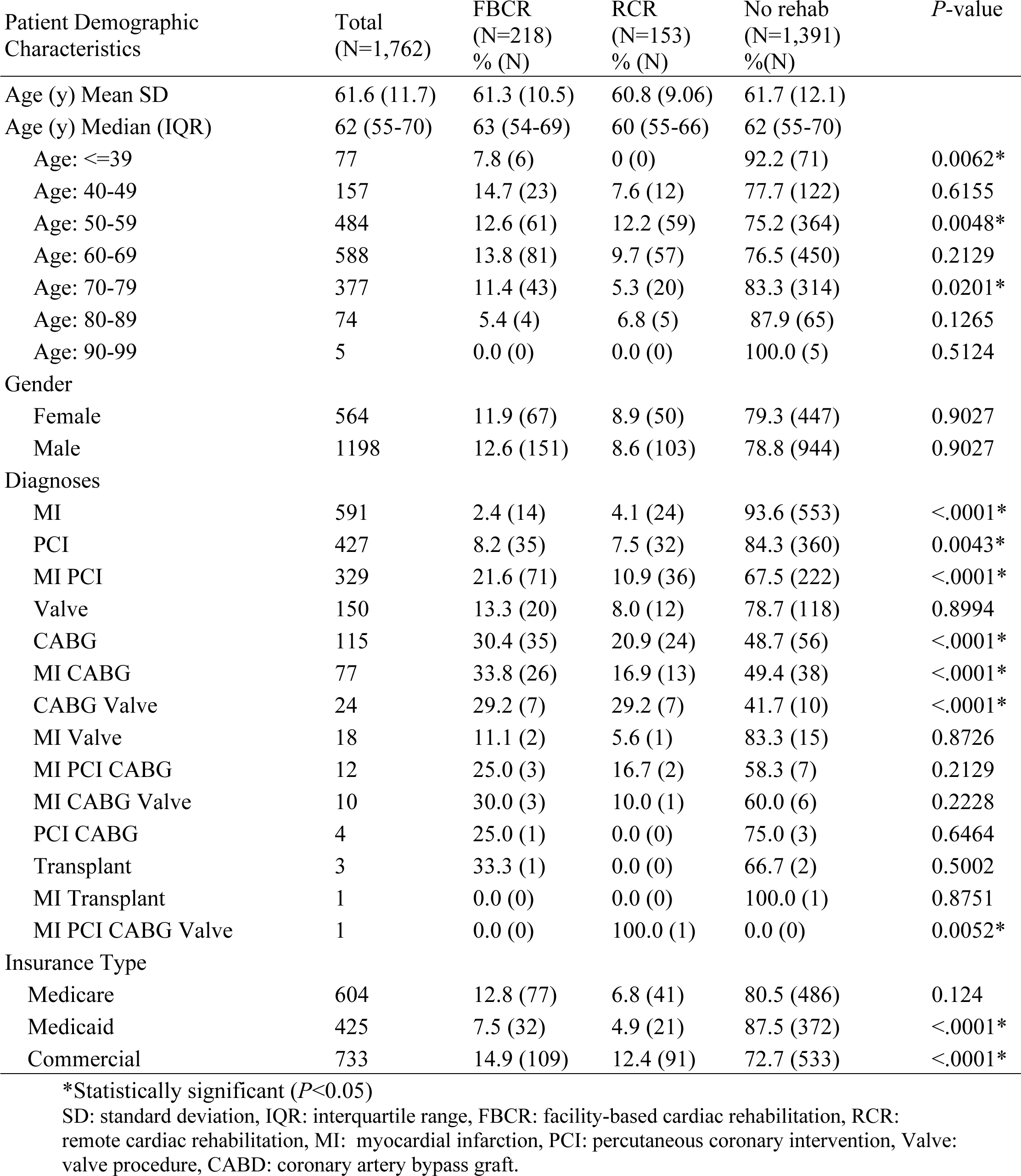
Patient Demographic Characteristics.

### Enrollment and Time to Enrollment

Data collection of the specific measures began in Q1 2021, one quarter before the intervention began; data prior to Q1 2021 were not available. Enrollment in FBCR went from 11.1% (32/286) to 16.2% (50/308). Enrollment in RCR went from .7% (2/286) (Q1, 2021) to 10.7% (33/308) (Q2, 2022) respectively. Total enrollment in CR increased from 11.9% (34/286) to 26.9% (83/308) (*P* <.0001) (see Figure 1). Note the RCR offering started in Q2; however, some patients who were discharged with qualifying diagnoses in Q1 2021 received the treatment in Q2 2021 and are captured in the data. Between Q1, 2021 and Q2, 2022 time to enrollment in FBCR went from 42 days (Q1 2021) to 47 days (Q2, 2022) (*P* = 0.1792) while time to enrollment for RCR decreased from 53 to 20 days (p < .0001), exceeding the recommended goal line of 21 days set by the AHA/ACC^37^. Overall, time to enrollment seemed to decrease by seven days by Q2, 2022 (p = 0.3348) (see Figure 2).

**Figure 1.**
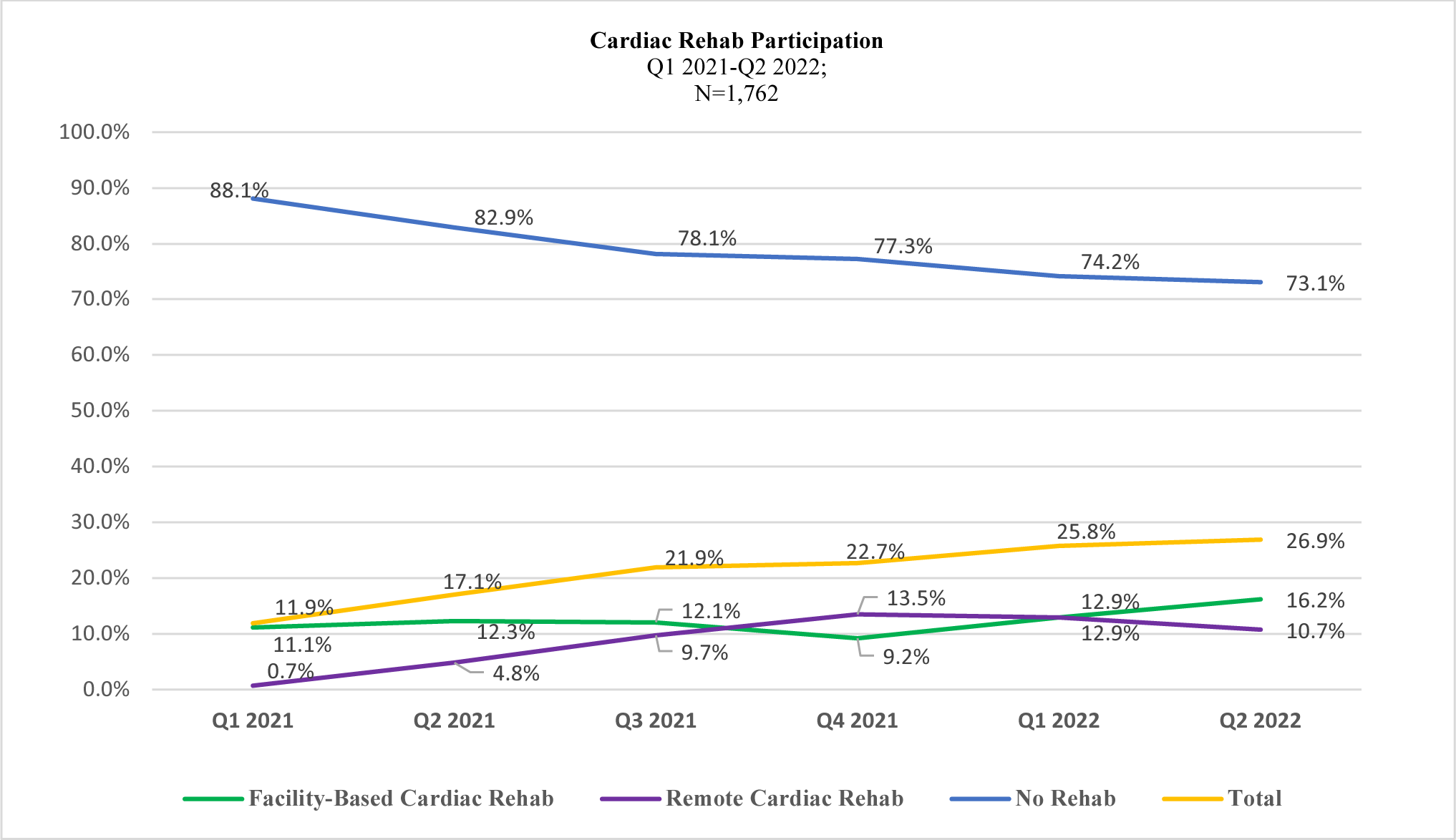

**Figure 2.**
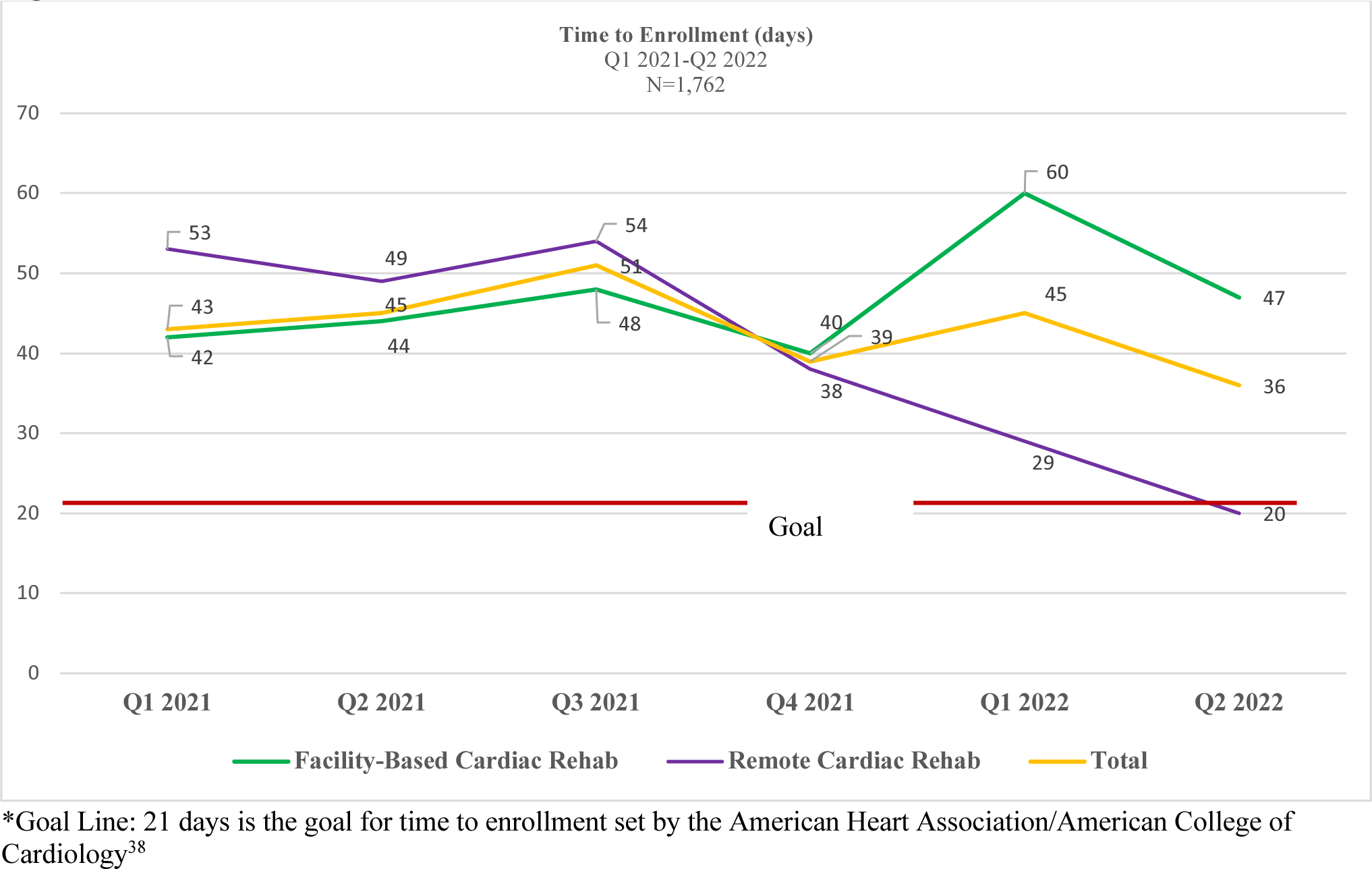

Overall, most patients did not enroll in CR (78.9%; 1391/1762). Compared to other groups, patients ≤ 39 years old enrolled less in CR and were more likely to attend FBCR over RCR if they attended (*P*=0.0062); patients between the 50-59 were more likely to attend RCR than FBCR (*P* =0.0048); and patients between the ages of 70-79, enrolled more in CR overall and attended FBCR over RCR (*P* =0.0201).

Patients with the diagnosis of MI only enrolled in CR less than other groups and this held true across FBCR and RCR (*P* <.0001); patients with PCI only looked very similar (*P* =0.0043); patients with MI followed by PCI enrolled in CR more than other groups and were more likely to attend FBCR than RCR (*P* <.0001); patients with CABG were more likely to get CR and attend FBCR more than RCR (*P* <0.0001); patients with MI CABG looked very similar as well (*P* <0.0001). Lastly, patients with CABG VALVE procedures were the most likely to enroll in CR with distribution of patients going to FBCR and RCR being fairly equal (*P* <.0001).

With regard to health insurance coverage, patients with commercial coverage were more likely to enroll in CR than other patients, and twice as likely to participate in RCR than other groups (*P* <.0001); patients with Medicaid coverage were less likely to enroll in CR overall and half as likely to attend FBCR or RCR than other groups (*P* <.0001).

## Discussion

The CRI was a concerted effort to improve access to CR by highlighting need, removing barriers, providing incentives, and introducing a RCR option for health plan members and providers. The goal was to promote uptake of CR. Within 15 months (April 2021-June 2022), overall participation in CR increased and time to enrollment improved, especially for RCR. Furthermore, we observed that RCR did not undermine FBCR participation, but rather complemented it. Patients with procedures clearly have a higher chance of being referred and thus enrolling in CR; and there is opportunity to improve CR for patients across all age groups and lines of business.

Further study to understand how and why performance in CR improved is warranted. Alternate options and the CRI focus overall could have also introduced competition forcing system change to accommodate patients as evidenced by the increase in CR overall. One could speculate that RCR improvement in time to enrollment may have been due to better coordination with local providers overtime given the new offering, or simply an increase in staffing which may be easier to accomplish in a virtual environment.

There were key learnings which, if addressed, could help to significantly amplify the initial successes of the initiative.

### Program Development

Program development to the launch of CRI took 18 months. To decrease this timeline, working in parallel to address cost share, physician incentives, credentialing and program implementation preparation as opposed to incrementally may accelerate the timeline to implementation. A staged approach where we rolled out RCR first while cost share was being addressed would have allowed a faster time to deployment (i.e.: 6 months). We learned that education of providers takes time and is an iterative process, thus, cost share waivers and/or physician incentives could have been introduced later as part of the overall rollout. Regardless, health plans who wish to replicate the CRI process need to be vigilant about timely filing of benefit waivers.

### Program Implementation

#### Initial Uptake of CRI

While received positively among the provider community, the CRI did not result in immediate increased enrollments in CR. This is reflected in the initial uptake of patients enrolling in RCR as well as the stable data related to enrollments in FBCR. Providers needed to be reminded of the benefits of CR, their performance on patient referrals, enrollments and adherence to CR, new available options (i.e., RCR) and what was in it for them (i.e., better patient outcomes, greater access and equity, physician incentives). Providing data, including benchmarking data, proved effective.

#### Acceptance of RCR

The gradual uptake of RCR was due to initial hesitation from providers. Part of this appeared to be due to system preferences to refer to FBCR within a health system. This was addressed by examining data and discussing delays in getting patients into FBCR in a timely window. Explaining that RCR maybe more suited to patients with low to moderate risk or without access to FBCR also validated the need for both FBCR and RCR. Lastly, referring patients to RCR could help with FBCR patient flow and intake especially if patients were transitioned from FBCR to RCR in a hybrid manner (i.e., patient starts off in FBCR and transitions to RCR). Concerns over the efficacy of RCR did come up; this was addressed by sharing data on effectiveness of RCR.

#### Generating Systematic Referrals to CR

Enrollments in RCR in the reported results were facilitated almost entirely by health plan care managers coordinating the referrals from available health plan data. In some cases, providers would coordinate with the health plan care managers to complete a referral. Since CDPHP was the only health plan with the CRI, improving the CR process for all patients seemed to be challenging for providers given the lack of consistency in offerings by insurance providers. Providers had to implement a different workflow for CDPHP members which was a rate limiting step to scaling referrals, especially referrals to RCR, which was not covered by other health plans.

One of the largest opportunities for increasing uptake of CR is accomplished with implementing automatic referrals for CR.^12,16^ Our query of providers revealed that some providers did have automatic referrals to CR while others lacked systematic processes for patient identification or referral. Our data reflected the fact that processes likely are in place for referring patients to CR post procedures, however, others such as those with MI and no procedure are likely not referred to CR. For hospitals who did have automated referrals, we learned that many were not including all eligible patients and were referring only to the system’s FBCR. In some cases, hospitals were eager to develop automated systems within their EMR however we encountered IT barriers preventing swift system change. EMR systems were often not built optimally and could not be amended in a timely fashion. We learned that it would take, in some cases, 1-2 years to accommodate a new process such as adding a RCR workflow.

#### Health Systems Change

The literature on practitioner behavior-change favors active multifaceted approaches, which we implemented (incentives, data feedback, one-on-one communication).^16, 39–40^ A key learning from this initiative is that effective systems change takes time. Providers need to understand the problem, digest the solution, and then implement change amongst a sea of competing health systems priorities.

Additionally, the strongest predictor of enrollment in CR is physician endorsement.^40–41^ Despite our concerted rollout efforts, we were unable to achieve consistent physician referral to CR in this early deployment phase. Moving the referral efforts upstream from the health plan liaisons to physicians and liaisons at the point of care is ideal. Studies of health systems that have combined automatic referrals to CR with education at the point of care have shown the greatest CR referral and subsequent enrollment rates (>86% referral, >74% enrollment).^27^

In the absence of CR policy mandates or public reporting and while quality metrics such as HEDIS CR metrics are still new, working with early adopter physician champions and administrative leaders who have the authority to shepherd efforts is critical.^26, 40–46^. Most of the recommendations in the Cardiac Rehabilitation Change Package are reliant on leadership to implement process change. Policy makers may want to consider policy mandates or public reporting to accelerate change as has been done for other facets of cardiac care.^43–46^

Even with leadership and potential reporting/policy change, as indicated in the Change Packet and referenced in the latest STEMI guidelines, a systems-based approach where CR providers keep patients engaged and the patient’s primary providers support completion is of utmost importance.^5, 13^ To this end, future endeavors to increase CR rates may consider a community collaboration with multiple stakeholders (i.e., hospitals, practices, CR providers, health plans, public health partners). In this effort, the CR stage can be set, goals and timeline for improvement defined, and a unified approach to addressing the problem can be tackled with physician leaders and administration included as key participants.

#### Study Limitations

The largest limitation of this project is that it was deployed in 2021 during the height of the COVID-19 pandemic. This limited the availably of true baseline data as there was little availability of FBCR prior to the implementation of the program. It may have also artificially inflated the number of patients who chose RCR over FBCR. Second, data constraints in terms of what was collected by the health plan prior to the study and available data such as full capture of race/ethnicity, limited our ability to conduct a true pre-post-comparison and more sophisticated analyses. Third, CDPHP incentivized physician practices and one could argue that hospitals and FBCR facilities also should have been incentivized. Fourth, access to referral data was not easily available and thus not included. Referral data may have been a better indicator of program adoption than enrollment and would have provided insight as to reasons why patients did not enroll after being referred. Fifth, our study was limited to one health plan with a relatively small size which could limit generalizability. Sixth, the multitude of interventions had different costs associated with them—parsing out the cost-effectiveness of each could not easily be assessed. Finally, given that we were only able to study 15 months of implementation (April 2021-June 2022), we reported on enrollment data and not participation or completion.

### Conclusion

The CRI introduced a paradigm shift and created momentum among providers to address modifiable barriers namely patient identification for CR and referral and enrollment processes. This study provides a structure that can be used to guide health plans to support referral, enrollment and participation in CR. The referenced frameworks to assess a community’s CR infrastructure and workflows may also be useful. Successful implementation of FBCR or RCR is dependent on clear referral and continued support processes for CR. Clinician and administrative leadership buy-in to CR is needed to create automatic systems and clinician endorsement for seamless referral and enrollment. Future efforts may see accelerated change with either a larger community wide initiative with multiple health plans and health system stakeholders at the table, or via policy mandates that support system change.

## Data Availability

The data used in this study belongs to The Capital District Physicians' Health Plan

## Acknowledgements

We thank Ian Brissette from the New York State Department of Health who supported this community effort from inception. We thank Michael Spicer and Misty Lundy from the Capital District Physicians’ Health Plan for their assistance on the details of the implementation and timelines. Additionally, we thank Thomas D’Aunno, Steven Levine and Ying Xian for their insight and thoughtful critique of this manuscript.

